# From Study Design to Executable Code: Automating Target Trial Emulation with Large Language Models

**DOI:** 10.64898/2026.03.13.26348306

**Authors:** Hanjae Kim, Minseong Kim, Seonji Kim, Seng Chan You

## Abstract

**Introduction:** Implementing target trial emulation (TTE) study methods as end-to-end executable analytic code is technically demanding, and producing standardized, reproducible scripts consistently across research teams remains a persistent challenge. We aimed to develop a framework that translates free-text study descriptions into standardized analytic specifications and executable Strategus R scripts for the Observational Health Data Sciences and Informatics (OHDSI) ecosystem.

**Methods:** We developed THESEUS (Text-guided Health-study Estimation and Specification Engine Using Strategus), which operates through two sequential steps. Large language models (LLMs) first map descriptions of the study into a constrained JavaScript Object Notation (JSON) schema (standardization step), after which the structured specifications are converted into R scripts with a self-auditing loop for error correction (code generation step). We evaluated eight proprietary LLMs using texts extracted from the methods section of 15 OHDSI-based TTE studies, and externally validated the framework on texts from 5 non-OHDSI studies, across three input settings: primary analysis text only, full analyses text, and full methods sections. Standardization was evaluated at the study-level (whether all parameters in a study were correctly extracted) and at the field-level (sensitivity and false positive rate per individual parameter) with field-level evaluation applied to the full analyses text and full methods sections input settings. Code generation was assessed by executability of the produced R scripts before and after self-auditing.

**Results:** In the standardization step, study-level accuracy across models ranged from 0.91 to 0.98 for primary analysis, 0.67 to 0.87 for full analyses, and 0.67 to 0.85 for full methods sections in OHDSI studies, whereas the corresponding ranges were 0.73 to 0.93, 0.60 to 0.87, and 0.27 to 0.47 in non-OHDSI studies. At the field-level, sensitivity across models under the full analyses text input setting ranged from 0.73 to 0.90 with 0.27 to 0.67 false positives per study in OHDSI studies, and from 0.71 to 0.90 with 0.20 to 1.00 false positives per study in non-OHDSI studies, depending on input setting. For code generation, first-run executability ranged from 0.80 to 1.00 for OHDSI studies and improved to 0.93 to 1.00 after self-auditing. In non-OHDSI studies, first-run executability ranged from 0.60 to 1.00, improving to 1.00 after self-auditing.

**Discussion:** THESEUS demonstrates that pairing a standardized data model with a structured analysis framework enables reliable LLM-powered automation of the coding step in observational research. THESEUS supports the reliable translation of natural-language study descriptions into executable, shareable code in standardized observational research settings. This approach has the potential to lower the technical barriers to participation in observational research for a broader range of investigators.

## INTRODUCTION

Comparative effectiveness research (CER) using observational data has become indispensable for generating real-world evidence on treatment effects, particularly when randomized controlled trials are infeasible due to ethical, logistical, or financial constraints. Target trial emulation (TTE), a methodological framework that explicitly mirrors the protocol of a hypothetical randomized trial to estimate causal effects from observational data,[1,2] has emerged as the standard approach for rigorous CER. Conducting a TTE study involves defining key components such as eligibility criteria, study periods, follow-up period with time zero, confounding control strategy, and outcomes,[1] each of which must be translated into executable code. This translation from a conceptual study design to a working analytic program remains a critical bottleneck, requiring both methodological expertise in causal inference and programming proficiency in statistical languages such as R or Python. Recent reviews have identified significant gaps in tool support for TTE, noting that no single tool currently supports all phases from design through analysis,[3] and that the primary barrier to broader adoption remains a lack of technical expertise rather than methodological understanding.[4]

The programming burden in CER is not merely a matter of inconvenience; it poses a fundamental barrier to the scalability and reproducibility of observational research. Each research team typically writes custom analytic code tailored to their local data environment, making it difficult to verify, replicate, or extend findings across institutions. Even when study designs are conceptually identical, differences in implementation such as variable naming, data transformations or even package versions can lead to divergent results. Addressing this challenge requires not only reducing the coding effort but also ensuring that the generated code operates on a standardized substrate where the same analytic logic can be faithfully executed across sites.

The Observational Health Data Sciences and Informatics (OHDSI)[5] community provides precisely such a substrate. OHDSI has built an international research network in which participating organizations convert their local data into the Observational Medical Outcomes Partnership Common Data Model (OMOP CDM), an open community data standard that harmonizes both the structure and the vocabulary of observational data across diverse sources.[6] Beyond data standardization, OHDSI has developed a comprehensive ecosystem of standardized open-source analytic tools.[7] The Health Analytics Data-to-Evidence Suite (HADES)[8] provides a collection of R packages for observational research, and Strategus[9] orchestrates these modules into fully reproducible workflows by generating a single JavaScript Object Notation (JSON) specification that encapsulates the entire analysis. A study coordinator can share this JSON file with collaborators at any OMOP CDM-converted site, who can then execute the analysis without writing or modifying any code. This combination of a standardized data model with a standardized analysis framework creates a uniquely favorable environment for automating the code generation process. Once a study design is formally specified, the mapping to executable code becomes deterministic rather than ad hoc. However, Strategus remains fundamentally an R package, and both HADES and Strategus require familiarity with OHDSI-specific conventions, which continues to limit accessibility for researchers who lack coding experience or come from other methodological traditions.

Recent advances in large language models (LLMs) have demonstrated their potential to bridge the gap between natural language and structured computations across various healthcare domains, including clinical trial design,[10] eligibility criteria conversion,[11,12] and electronic health record phenotyping.[13] We hypothesized that the standardized nature of OMOP CDM, combined with the well-defined structure of Strategus analysis specifications, would make this ecosystem particularly amenable to LLM-powered code generation because the target output format is constrained and predictable rather than open-ended. Inspired by prior work on SOCRATex,[14] a hierarchical standardization framework that transforms free-text clinical narratives into structured, machine-readable representations, we aimed to develop a process which adopts a two-step approach consisting of standardization and code generation to systematically translate study designs into Strategus codes. The developed framework, THESEUS (Text-guided Health-study Estimation and Specification Engine Using Strategus), standardizes natural language study descriptions into structured JSON-based analytic specifications and then converts these specifications into executable Strategus R scripts. THESEUS aims to make TTE more accessible to a broader range of researchers by lowering the learning curve for those entering the OHDSI ecosystem, and to enhance the reproducibility of observational studies conducted across the OHDSI network.

## METHODS

We report the details of this study in accordance with the Minimum Reporting Items for Clear Evaluation of Accuracy Reports of Large Language Models in Healthcare (MI-CLEAR-LLM) checklist.[15]

### Conceptual framework design

As proof-of-concept, we focused on implementing ‘Cohort Method’ design within the OHDSI research framework which emulates randomized controlled trials. This requires specifying the study period, time-at-risk (TAR) defined as follow-up window, and propensity score (PS) adjustment strategy for confounding control. We adopted a two-step approach consisting of standardization and code generation to reflect natural language study descriptions into Strategus-compatible analytic code (Figure 1).

**Figure 1.**
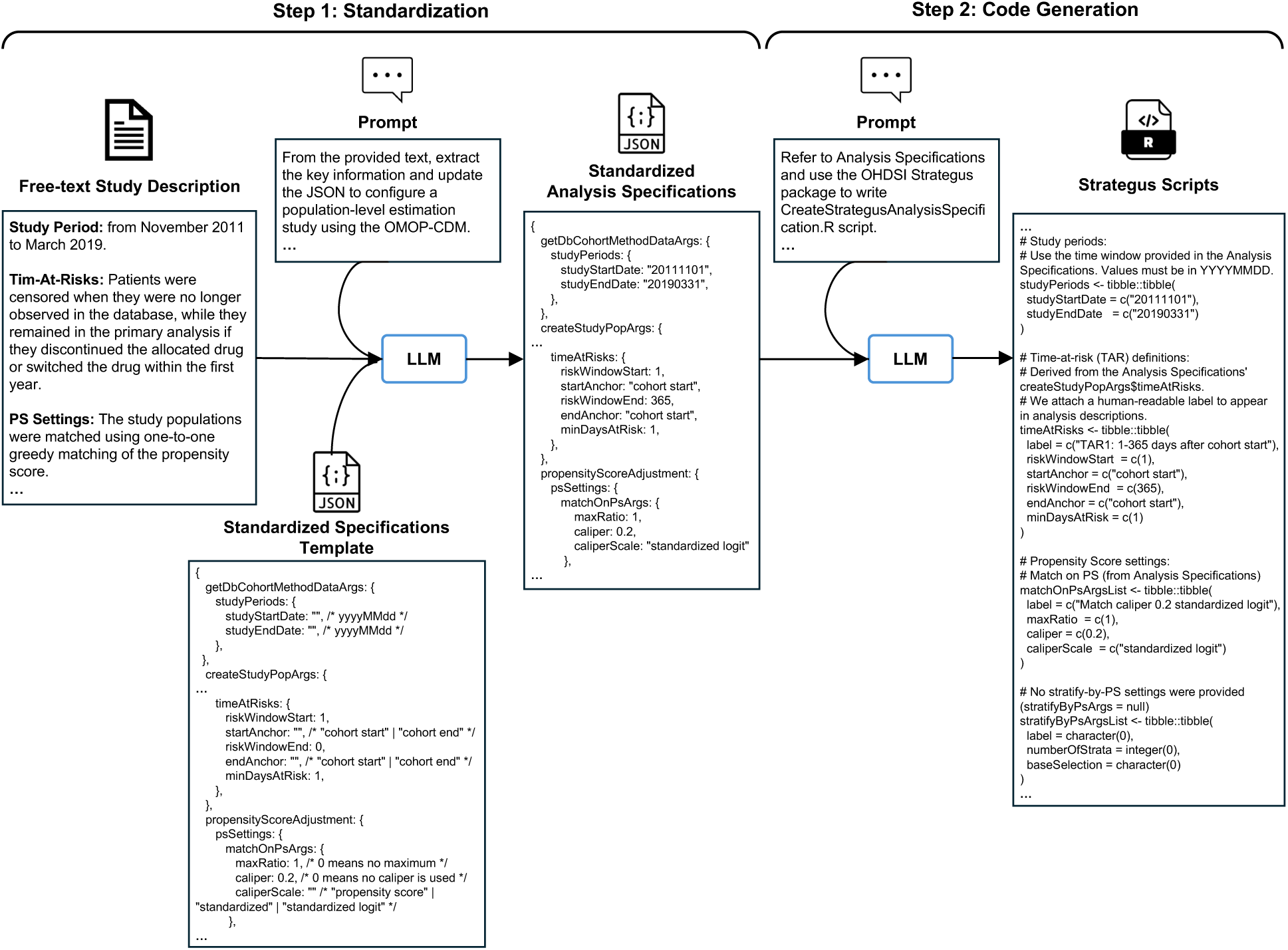
Conceptual framework of THESEUS for translating free-text study descriptions into Strategus scripts. LLM indicates large language model

### Step 1: Standardizing free-text study descriptions into structured specifications

When a researcher plans a study in natural language, this needs to be translated into a standardized OHDSI research framework. We leveraged a JSON-based schema that is commonly used within the OHDSI ecosystem to represent study design components. This schema encompasses all items required to specify an observational study. In this study, we restricted its scope to the fields related to the study period, TAR, and PS adjustment (Supplementary Material 1). An LLM takes text descriptions of study designs together with the schema template as inputs and generates a structured specification in JSON format. In addition, the LLM produces an explanation describing how each part of the text was interpreted and applied to the analytic specifications. To facilitate accurate interpretation of the study design context, we added descriptions of each field of the schema, referenced from the OHDSI guideline book,[5] directly into the prompt. The prompt used for this standardization step is provided in Supplementary Material 2.

### Step 2: Generating Strategus R scripts based on structured study specifications

The structured specifications generated in the previous step are then transformed into executable Strategus R scripts. Based on the specifications, the LLM generates Strategus R scripts with embedded annotations that provide human-readable explanations for each code line. The prompt includes a predefined template of Strategus scripts to ensure correctness and consistent structure (Supplementary Material 3). In addition, we incorporated a self-auditing function in this step, in which the LLM can review and correct the generated script when execution errors are detected. The resulting scripts can be directly copied into the ‘CreateStrategusAnalysisSpecification.R’ file of the Strategus study repository template cloned from GitHub[16] and executed without modification. Upon execution, this R script generates the final JSON file required to run the actual analysis, which can then be shared directly with collaborating researchers. The prompts used for code generation and self-auditing in this step are provided in Supplementary Material 4 and 5.

### Development of a graphical user interface prototype

The OHDSI community provides a web-based platform called ATLAS,[17] which provides an environment for configuring various analysis specifications within a unified graphical user interface (GUI). To enable human-in-the-loop validation of the standardized specifications before conducting the code generation step, we developed a GUI prototype that resembles the ‘population-level estimation’ tab of ATLAS version 2.14.1 (Supplementary Material 6), incorporating the proposed agentic framework. Similar to the original ATLAS interface, users can configure analysis settings with manual clicks, while they can also input free-text study descriptions, which will be translated into corresponding analysis specifications displayed on the GUI. Before updating the GUI, the system presents a side-by-side comparison of the original and revised specifications, allowing users to selectively accept or reject individual modifications. Once the configuration is finalized, a single click of a button enables conversion of the GUI-based analysis specifications into executable Strategus R scripts. Figure 2 illustrates the overall workflow through the GUI prototype.

**Figure 2.**
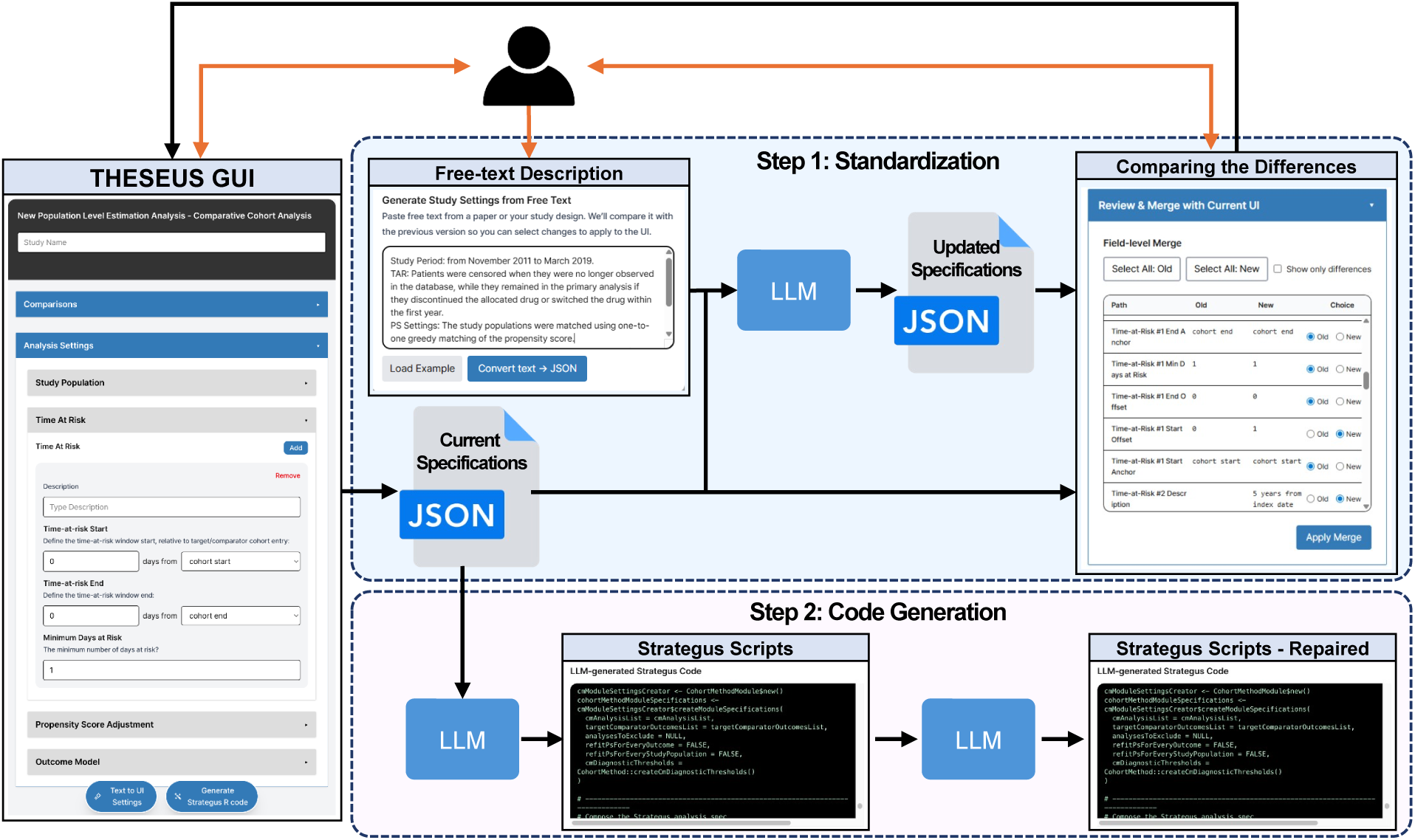
Overall workflow of THESEUS powered by LLMs with human-in-the-loop configuration The orange colored-arrows represent the interaction between human and the system.

### Evaluation of the framework

Previously published TTE study papers[18–32] that used OMOP CDM and OHDSI R packages (either HADES or Strategus) were used as natural language samples to evaluate the framework’s ability to standardize free-text study descriptions into structured analytic specifications. All texts from the papers were manually extracted by the authors. We initially used only the text describing the study period, TAR, and PS adjustment for the primary analysis extracted from each study as LLM inputs. Because TTE studies commonly include sensitivity analyses that compare partially modified study designs, we further evaluated the LLMs under two additional input conditions of multiple analyses settings: (1) providing all text related to all analyses (full analyses), including both primary and sensitivity analyses, but limited to descriptions of the study period, TAR, and PS adjustment; and (2) providing the full methods section text from each paper. These two input conditions allowed the generated JSON specifications to include multiple fields within each section of the schema (e.g., multiple risk windows within the TAR section for a single study). Across all three input settings, each study was processed using independent LLM inferences, and the resulting structured specifications were compared with gold standards created by the authors. Accuracy was assessed separately for each of the three specification sections (study-level evaluation). For the multiple analyses settings (full analyses and full methods section), where a single section could contain multiple fields, each section was counted as correct only when all corresponding subfields exactly matched the gold standards. Only for the multiple analyses input settings, we additionally performed field-level evaluation, treating each subfield as an independent unit and aggregating them across all studies to compute sensitivity, precision, F1-scores, and the number of false positives (FPs) per study.

For evaluation of the code generation step, the JSON specifications reviewed and finalized by the authors during the standardization step evaluation were used as inputs under both the primary analysis and multiple analyses settings. The scripts generated by the LLM during both the initial generation and the subsequent self-auditing step were reviewed and executed to verify that they ran without errors. All script executions in this study were performed using R version 4.5.1 with Strategus version 1.4.1 and Java version 17.0.17.

For all evaluations, we compared a diverse set of proprietary LLMs from 4 major vendors: OpenAI (GPT-5.2, GPT-4.1), Google (Gemini-2.5-Pro, Gemini-2.5-Flash), Anthropic (Claude-Opus-4.5, Claude-Haiku-4.5), and DeepSeek (DeepSeek-V3.2 Thinking Mode, DeepSeek-V3.2 Non-thinking Mode). Detailed model versions are specified in Supplementary Material 7. All models were accessed via their application programming interfaces (APIs) to fix model versions at the time of experimentation and to avoid inter-session memory accumulation. No model fine-tuning was performed in this study. All model queries were executed in January 2026. To minimize variability across inference attempts, the temperature parameter was set to 0. Each input prompt was executed as a single attempt without retries. The same logical prompt content with no system messages and no model-specific prompt modification was passed across all vendors.

To assess the generalizability of the framework beyond the OHDSI community, we additionally conducted an external validation using non-OHDSI study papers.[33–37] We included TTE studies that explicitly described the study period, TAR, and PS adjustment. Studies that employed inverse probability weighting (IPW) for PS adjustment were excluded, as this approach fell outside the scope of the current system design. All included non-OHDSI studies underwent the same evaluation procedures as the OHDSI-based studies.

## RESULTS

### Resulting Prototype and Public Resources

An interactive web-based prototype is available at https://theseus2.vercel.app/. A demonstration video of the prototype is available at https://youtu.be/xUWI4e-78Wk.

### Standardization Evaluation Results

At the study-level, across the 15 OHDSI-based studies, overall accuracy under the primary analysis input condition ranged from 0.91 to 0.98 across models, with Claude-Opus-4.5 achieving the highest (overall 0.98; study period 1.00, TAR 0.93, PS adjustment 1.00) (Table 1). Under the full analyses input condition, overall accuracy ranged from 0.67 to 0.87, with Claude-Opus-4.5 again achieving the highest (overall 0.87; study period 1.00, TAR 0.73, PS adjustment 0.87). When the full methods section text was provided as input, overall accuracy ranged from 0.67 to 0.85, with DeepSeek-V3.2 Non-thinking Mode achieving the highest (overall 0.85; study period 0.87, TAR 1.00, PS adjustment 1.00).

**Table 1.** Study-level standardization accuracy across OHDSI and non-OHDSI studies under three input conditions. OHDSI indicates Observational Health Data Sciences and Informatics; SP, Study Period; TAR, Time-at-Risk; PS, Propensity Score (adjustment).

| OHDSI studies (N=15) |  |  |  |  |  |  |  |  |  |  |  |  |
| --- | --- | --- | --- | --- | --- | --- | --- | --- | --- | --- | --- | --- |
| Models | Accuracy |  |  |  |  |  |  |  |  |  |  |  |
|  | Primary analysis only |  |  |  | Full analyses |  |  |  | Full methods section |  |  |  |
|  | SP | TAR | PS | Overall | SP | TAR | PS | Overall | SP | TAR | PS | Overall |
| GPT-5.2 | 1.0 | 0.80 | 0.93 | 0.91 | 0.93 | 0.80 | 0.80 | 0.84 | 0.73 | 0.73 | 0.87 | 0.78 |
| GPT-4.1 | 1.0 | 0.80 | 1.0 | 0.93 | 0.87 | 0.33 | 0.80 | 0.67 | 0.67 | 0.60 | 0.73 | 0.67 |
| Gemini-2.5-Pro | 1.0 | 0.80 | 1.0 | 0.93 | 0.87 | 0.67 | 0.80 | 0.78 | 0.73 | 0.67 | 0.73 | 0.71 |
| Gemini-2.5-Flash | 1.0 | 0.87 | 1.0 | 0.96 | 0.93 | 0.73 | 0.67 | 0.78 | 0.80 | 0.67 | 0.67 | 0.71 |
| Claude-opus-4.5 | 1.0 | 0.93 | 1.0 | 0.98 | 1.0 | 0.73 | 0.87 | 0.87 | 0.73 | 0.80 | 0.93 | 0.82 |
| Claude-haiku-4.5 | 1.0 | 0.73 | 1.0 | 0.91 | 1.0 | 0.60 | 0.73 | 0.78 | 0.87 | 0.53 | 0.87 | 0.76 |
| DeepSeek-V3.2 Thinking Mode | 1.0 | 0.80 | 1.0 | 0.93 | 0.93 | 0.73 | 0.87 | 0.84 | 0.87 | 0.67 | 0.93 | 0.82 |
| DeepSeek-V3.2 Non- Thinking Mode | 1.0 | 0.80 | 1.0 | 0.93 | 0.87 | 0.60 | 0.80 | 0.76 | 0.87 | 0.67 | 1.0 | 0.85 |
| Non-OHDSI studies (N=5) |  |  |  |  |  |  |  |  |  |  |  |  |
| Models | Accuracy |  |  |  |  |  |  |  |  |  |  |  |
|  | Primary analysis only |  |  |  | Full analyses |  |  |  | Full methods section |  |  |  |
|  | SP | TAR | PS | Overall | SP | TAR | PS | Overall | SP | TAR | PS | Overall |
| GPT-5.2 | 1.0 | 0.40 | 0.80 | 0.73 | 0.80 | 0.60 | 1.0 | 0.80 | 0.60 | 0.20 | 0.60 | 0.47 |
| GPT-4.1 | 1.0 | 0.40 | 1.0 | 0.80 | 0.80 | 0.20 | 0.80 | 0.60 | 0.60 | 0.0 | 0.20 | 0.27 |
| Gemini-2.5-Pro | 1.0 | 0.80 | 0.80 | 0.87 | 1.0 | 0.60 | 0.80 | 0.80 | 0.60 | 0.0 | 0.40 | 0.33 |
| Gemini-2.5-Flash | 1.0 | 0.80 | 0.80 | 0.87 | 0.80 | 0.80 | 0.80 | 0.80 | 0.80 | 0.0 | 0.40 | 0.40 |
| Claude-opus-4.5 | 1.0 | 0.80 | 1.0 | 0.93 | 1.0 | 0.60 | 1.0 | 0.87 | 0.60 | 0.40 | 0.40 | 0.47 |
| Claude-haiku-4.5 | 1.0 | 0.40 | 0.80 | 0.73 | 1.0 | 0.60 | 0.80 | 0.80 | 0.80 | 0.40 | 0.20 | 0.47 |
| DeepSeek-V3.2 Thinking Mode | 1.0 | 1.0 | 0.80 | 0.93 | 0.80 | 0.80 | 1.0 | 0.87 | 0.60 | 0.0 | 0.40 | 0.33 |
| DeepSeek-V3.2 Non- Thinking Mode | 1.0 | 0.80 | 0.80 | 0.87 | 0.80 | 0.60 | 0.80 | 0.73 | 0.60 | 0.40 | 0.20 | 0.40 |

Across the 5 non-OHDSI studies, overall accuracy under the primary analysis input condition ranged from 0.73 to 0.93, with Claude-Opus-4.5 and DeepSeek-V3.2 Thinking Mode jointly achieving the highest overall accuracy of 0.93 (Table 1). Under full analyses input condition, overall accuracy ranged from 0.60 to 0.87, with Claude-Opus-4.5 and DeepSeek-V3.2 Thinking Mode again achieving the highest (overall 0.87). When the full methods section text was provided as input, overall accuracy decreased, ranging from 0.27 to 0.47, with GPT-5.2, Claude-Opus-4.5, and Claude-Haiku-4.5 achieving the highest (overall 0.47).

At the field-level evaluation for multiple analyses input conditions, the 15 OHDSI studies yielded 84 specification items (study period 18, TAR 35, PS adjustment 31). Under full analyses input condition, precision ranged from 0.88 to 0.95, sensitivity from 0.73 to 0.90, with false positives per study ranging from 0.27 to 0.67 across all models (Table 2). DeepSeek-V3.2 Thinking Mode achieved the strongest field-level performance (precision 0.95, sensitivity 0.89, false positives per study 0.27). When the full methods section text was provided as input, precision ranged from 0.88 to 0.95, sensitivity from 0.76 to 0.89, with false positives per study ranging from 0.27 to 0.60; DeepSeek-V3.2 Non-thinking Mode achieved the highest precision (0.95) with the lowest false positives per study (0.27).

**Table 2.** Field-level precision and sensitivity on gold-standard specification items. FP indicates false positive; SP, Study Period; TAR, Time-at-Risk; PS, Propensity Score (adjustment).

| **OHDSI studies (N=15)** |  |  |  |  |  |  |
| --- | --- | --- | --- | --- | --- | --- |
| **Models** | **Full analyses (n=84)** |  |  | **Full methods section (n=84)** |  |  |
|  | Precision | Sensitivity | FP per study | Precision | Sensitivity | FP per study |
| GPT-5.2 | 0.91 | 0.73 | 0.40 | 0.94 | 0.76 | **0.27** |
| GPT-4.1 | 0.94 | **0.90** | 0.33 | 0.94 | 0.87 | 0.33 |
| Gemini-2.5-Pro | 0.88 | 0.86 | 0.67 | 0.88 | 0.82 | 0.60 |
| Gemini-2.5-Flash | 0.91 | 0.86 | 0.47 | 0.90 | 0.82 | 0.53 |
| Claude-opus-4.5 | 0.91 | 0.89 | 0.47 | 0.90 | 0.88 | 0.53 |
| Claude-haiku-4.5 | 0.90 | 0.86 | 0.53 | 0.89 | 0.83 | 0.43 |
| DeepSeek-V3.2 Thinking Mode | **0.95** | 0.89 | **0.27** | 0.93 | **0.89** | 0.40 |
| DeepSeek-V3.2 Non-thinking Mode | 0.92 | 0.83 | 0.40 | **0.95** | 0.88 | **0.27** |
| **Non-OHDSI studies (N=5)** |  |  |  |  |  |  |
| **Models** | **Full analyses (n=21)** |  |  | **Full methods section (n=21)** |  |  |
|  | Precision | Sensitivity | FP per study | Precision | Sensitivity | FP per study |
| GPT-5.2 | 0.90 | 0.86 | 0.40 | **0.50** | 0.48 | **2.0** |
| GPT-4.1 | 0.75 | 0.71 | 1.0 | 0.36 | 0.43 | 3.2 |
| Gemini-2.5-Pro | 0.86 | 0.86 | 0.60 | 0.40 | 0.48 | 3.0 |
| Gemini-2.5-Flash | 0.90 | 0.86 | 0.40 | 0.42 | **0.52** | 3.0 |
| Claude-opus-4.5 | 0.90 | **0.90** | 0.40 | 0.38 | 0.48 | 3.2 |
| Claude-haiku-4.5 | 0.86 | 0.86 | 0.60 | 0.44 | **0.52** | 2.8 |
| DeepSeek-V3.2 Thinking Mode | **0.95** | **0.90** | **0.20** | 0.42 | 0.38 | 2.2 |
| DeepSeek-V3.2 Non-thinking Mode | 0.85 | 0.81 | 0.60 | **0.50** | 0.48 | **2.0** |

For the 5 non-OHDSI studies, 21 specification items were evaluated (study period 6, TAR 6, PS adjustment 9). Under full analyses input condition, precision ranged from 0.75 to 0.95, sensitivity from 0.71 to 0.90, with false positives per study ranging from 0.20 to 1.0 (Table 2). DeepSeek-V3.2 Thinking Mode achieved the strongest field-level performance (precision 0.95, sensitivity 0.90, false positives per study 0.20). When the full methods section text was provided as input, precision ranged from 0.36 to 0.50, sensitivity from 0.38 to 0.52, with false positives per study ranging from 2.0 to 3.2.

### Code Generation Evaluation Results

For code generation, executability was defined as the proportion of studies for which the generated Strategus script executed without errors. In the 15 OHDSI studies under the primary analysis condition, first-run executability ranged from 0.80 to 1.00, with GPT-5.2, GPT-4.1, and Claude-Opus-4.5 achieving 1.00. After self-auditing, executability increased to 0.93-1.00, with most models achieving 1.00 (Figure 3). Under multiple analyses condition, first-run executability ranged from 0.67 to 1.00; GPT-5.2, GPT-4.1, Claude-Opus-4.5 achieving the highest executability. After self-auditing, executability improved across all models (0.93–1.00), with multiple models reaching 1.00.

**Figure 3.**
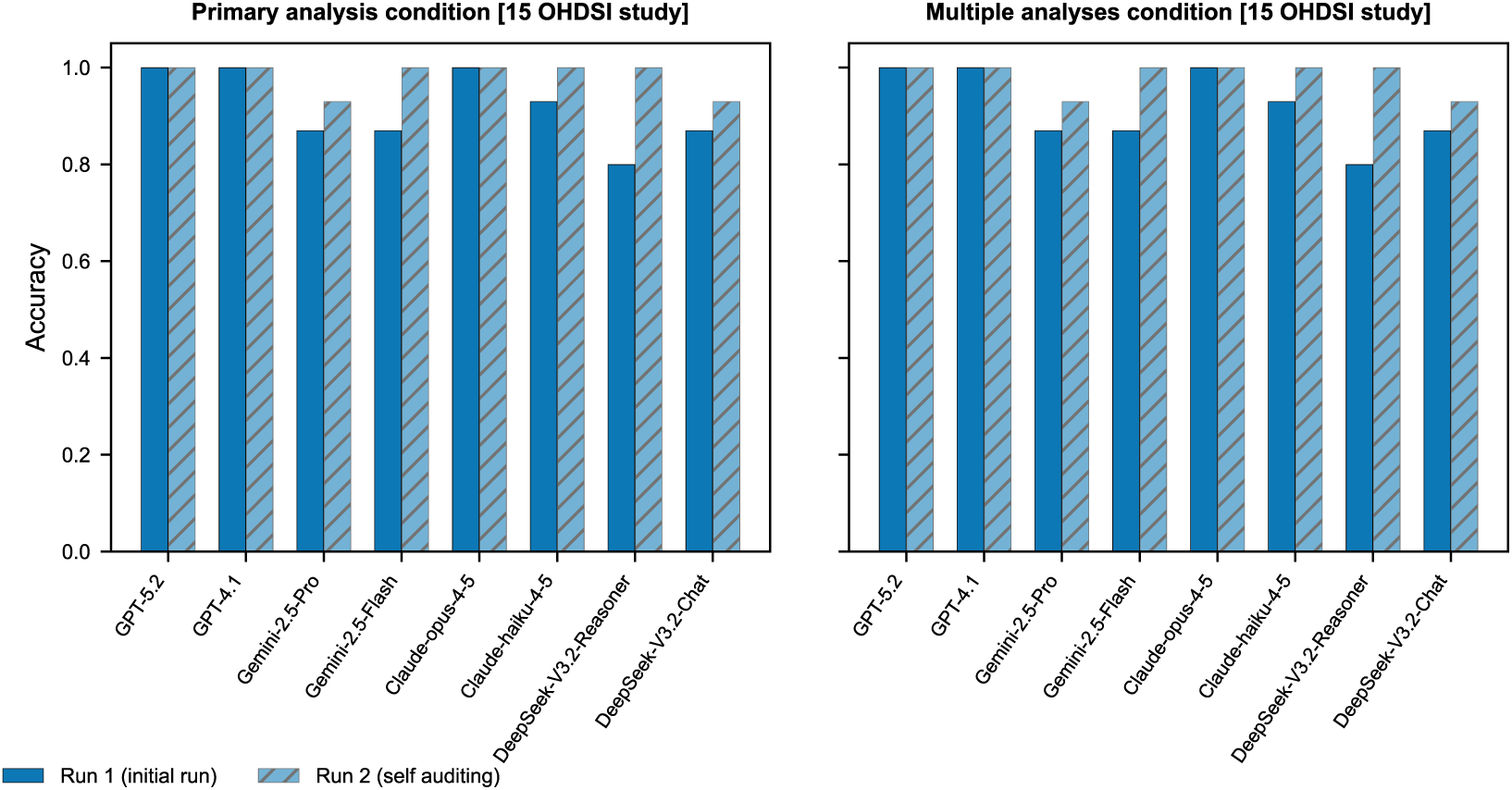
Code-generation executability across 15 OHDSI studies before and after self-auditing. Bars show the proportion of studies in which the generated Strategus script executed without errors (Run 1) and after a self-auditing revision (Run 2), under Primary analysis only and Full analyses conditions.

In the 5 non-OHDSI studies, under the primary analysis condition, first-run executability was 1.00 for most models; Gemini-2.5-Flash (0.80), Claude-Opus-4.5 (0.80), and DeepSeek-V3.2 Thinking Mode (0.60) were below 1.00. After self-auditing, all models achieved 1.00. Under multiple analyses condition, first-run executability ranged from 0.80 to 1.00, and all models achieved 1.00 after self-auditing (Figure 4).

**Figure 4.**
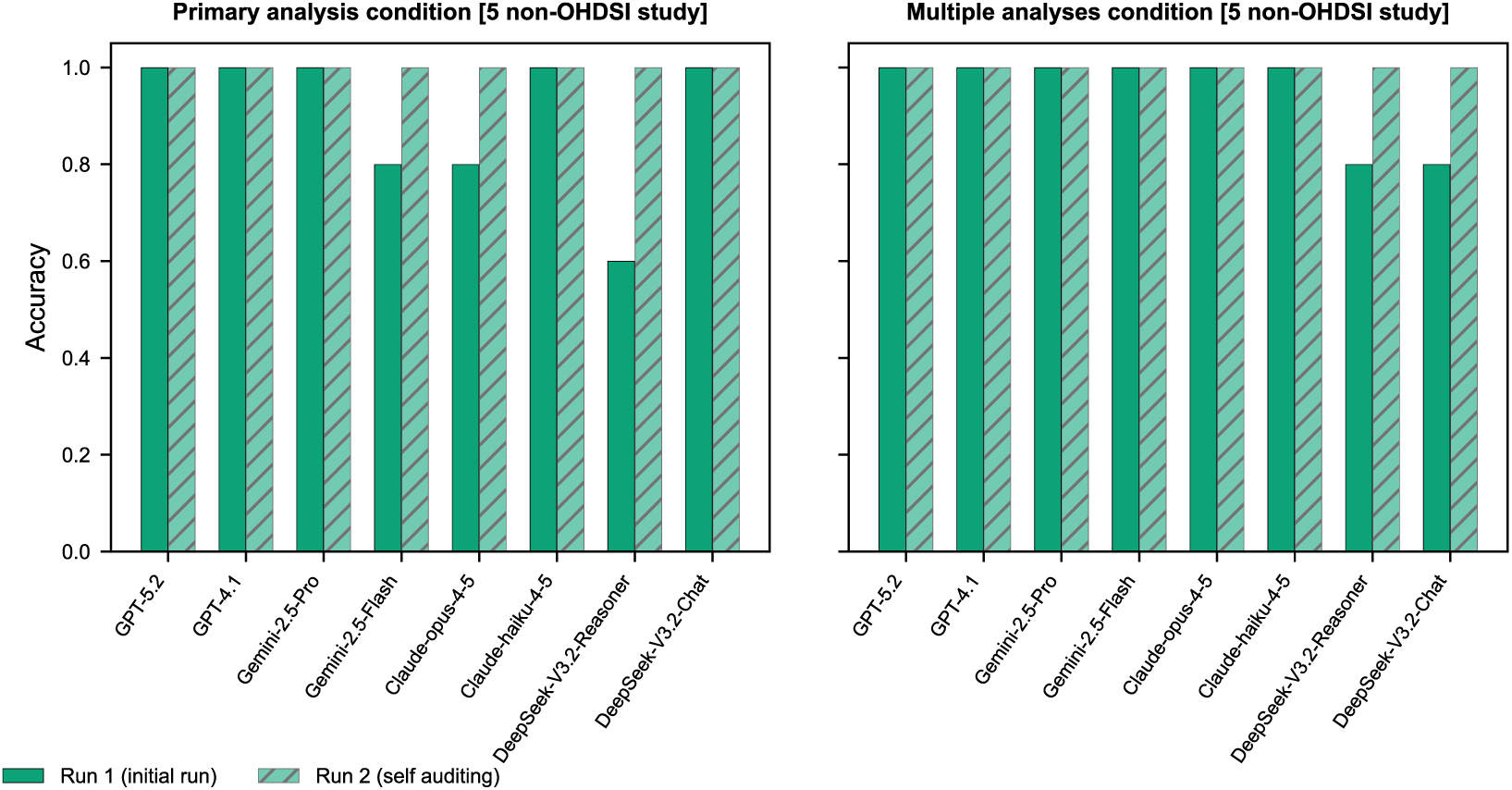
Code-generation executability across 5 non-OHDSI studies before and after self-auditing. Bars show the proportion of studies in which the generated Strategus script executed without errors (Run 1) and after a self-auditing revision (Run 2), under Primary analysis and Full analyses conditions.

## DISCUSSION

This study demonstrates that the coding step of CER can be substantially automated when the underlying data and analytic infrastructure are standardized. By leveraging the structured nature of OMOP CDM and the well-defined specification format of Strategus, THESEUS translates natural language study descriptions into executable analytic code through a two-step framework of standardization and code generation while preserving human oversight. Our results show that current LLMs can extract core analytic parameters from natural language study descriptions with high accuracy and convert structured specifications into reliable, executable R scripts, thereby bridging the gap between how researchers conceptualize a study and how it is computationally implemented.

A key insight from this work is that the success of LLM-powered code generation critically depends on the degree of standardization in both the data layer and the analytic layer. Even when observational studies address the same research question, independent research teams may implement the study design differently, leading to heterogeneous results that reduce the reproducibility of the studies.[38,39] OHDSI’s ecosystem resolves this problem through two layers of standardization: OMOP CDM provides a universal data structure and vocabulary, while Strategus provides a universal specification format for analyses. Together, these layers constrain the code generation task from an open-ended programming problem into a structured mapping problem, where the input (a JSON specification) and the output (an R script) both conform to well-defined schemas. This is what makes the OHDSI ecosystem uniquely amenable to automation, and we believe other standardized research platforms in diverse domains could similarly benefit from LLM-assisted code generation.

Our evaluation of the standardization step showed that LLMs have strong potential to extract core analytic settings from natural-language descriptions, though performance varied across specification components. Study period and propensity score adjustment specifications were extracted with consistently high accuracy across models, while time-at-risk extraction proved more challenging, likely reflecting the greater ambiguity and diversity in how this parameter is described in published methods sections. When sensitivity analyses were included, overall field-level performance showed high sensitivity with low false positive rates (less than one per study, except when full methods section text was provided for non-OHDSI studies). The external validation with non-OHDSI studies further demonstrates that the standardization step can generalize beyond the OHDSI community, enabling retrospective reconstruction of analytic specifications from studies conducted in different data environments.

The code generation step demonstrated that LLMs can generate Strategus R scripts with high reliability. By design, this step takes structured, machine-readable specifications as input, which organize the numerous configuration parameters required by Strategus into a systematic format, enabling consistent conversion into executable code. This intermediate representation also decouples the code generation from the complexities of natural language interpretation, allowing separate validation of the study design before execution. The inclusion of a self-auditing step substantially improved execution success rates, with multiple models achieving perfect execution after self-correction. This finding underscores a broader principle for LLM-based code generation: combining structured input representations with iterative self-correction can yield near-deterministic reliability even for complex analytic pipelines.

Cohort definition remains one of the most complex and labor-intensive steps in observational studies,[40] and complementary efforts are advancing its automation. Park et al. developed Criteria2Query 3.0,[12] a system that transforms free-text clinical trial eligibility criteria into executable OMOP CDM–compatible queries using LLMs for concept extraction and Structured Query Language (SQL) generation. More recently, Lee et al. proposed a similar framework with systematic evaluation across multiple LLMs.[11] Integrating such cohort definition pipelines with THESEUS could enable end-to-end automation of the entire TTE workflow, from defining study populations to generating executable analysis code. Furthermore, as demonstrated by our external validation, such an integrated system could also facilitate retrospective verification of previously published studies by reconstructing their analytic specifications from reported methods.

This study has several limitations. First, the current prototype is limited to TTE framework. However, the modular architecture of THESEUS can be expanded to support a broader range of study designs such as characterization or patient-level prediction. Second, our framework currently supports only matching and stratification for PS adjustment, as these are the predominant approaches within the OHDSI ecosystem.[5] Consequently, studies employing IPW were excluded from evaluation, which may have introduced selection bias given the widespread use of this method in the broader TTE literature. Third, the sample size used for evaluation was small, as constructing gold-standard specifications from published study papers is labor-intensive and requires domain expertise. As an initial feasibility study, our findings provide preliminary evidence of the framework’s potential, but larger-scale evaluations will be necessary to establish generalizability. Fourth, the framework’s reliance on OHDSI-specific tools means that researchers at institutions without OMOP CDM-converted databases would need to undergo data conversion before benefiting from automated code generation – though the growing global adoption of OMOP CDM is progressively lowering this barrier.

In conclusion, THESEUS demonstrates that the combination of standardized data models and standardized analysis frameworks creates a fertile ground for LLM-powered automation of the coding step in CER. This work addresses a key bottleneck that has limited the scalability and accessibility of observational studies by enabling researchers to express study designs in natural language. As standardized data networks continue to expand globally, the principles of leveraging structural constraints to enable reliable automation may generalize to other research ecosystems and study types, ultimately broadening participation in evidence generation from real-world data.

## Data Availability

The source code for the application is available at https://github.com/dr-you-group/theseus-app.

## Author Contributions

**Conceptualization**: HK, SCY. **Data curation**: HK, MSK. **Formal analysis**: HK, MSK. **Funding acquisition**: SCY. **Investigation**: HK, MSK, SK. **Methodology**: HK, MSK, SCY. **Project administration**: HK, SCY. **Resources**: HK, MSK, SCY. **Software**: MSK. **Supervision**: SCY. **Validation**: HK, MSK, SK. **Visualization**: HK, MSK. **Writing – original draft**: HK, MSK. **Writing – review & editing**: HK, MSK, SK, SCY.

---

**Supplementary Material 1.**
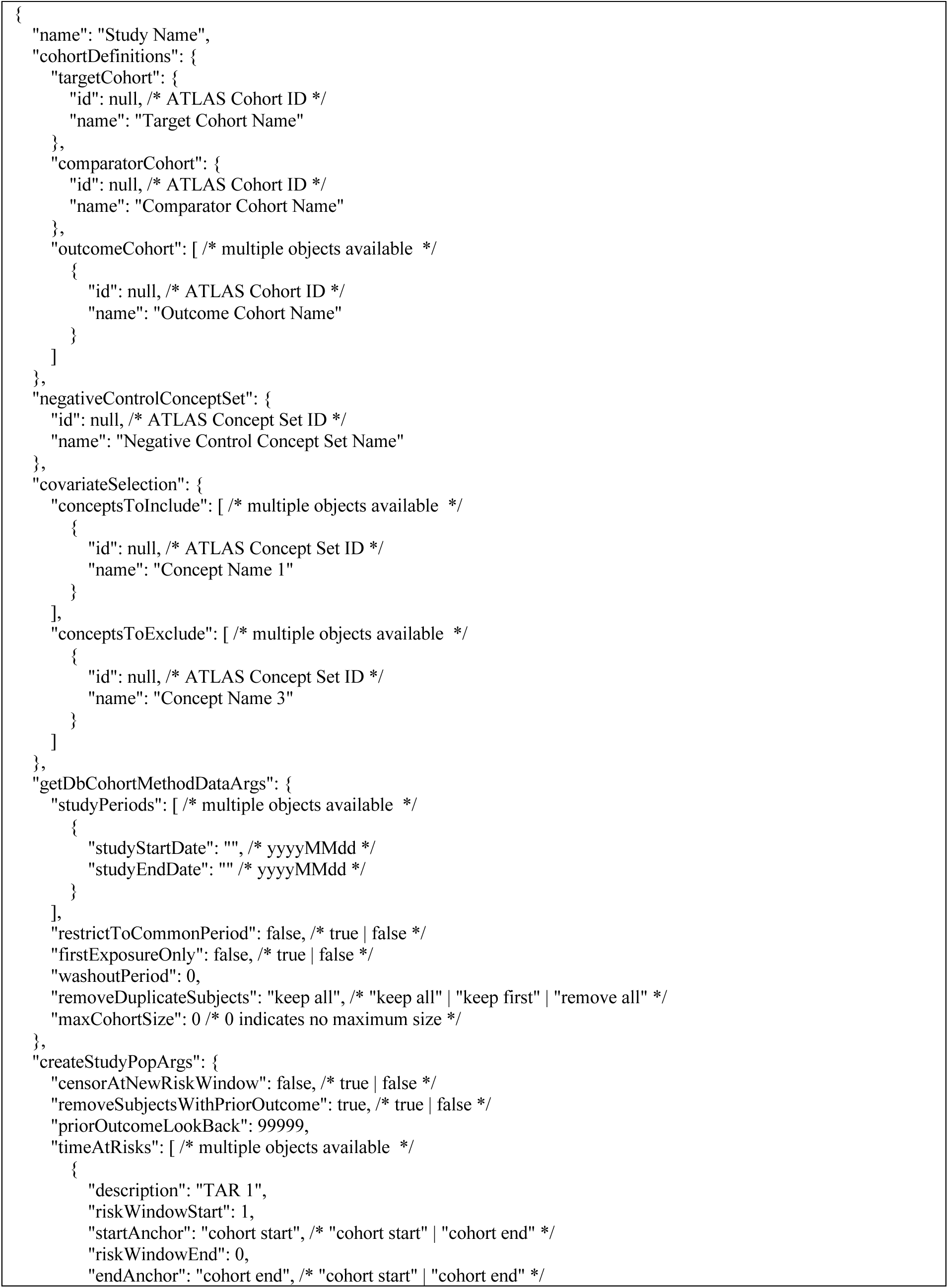
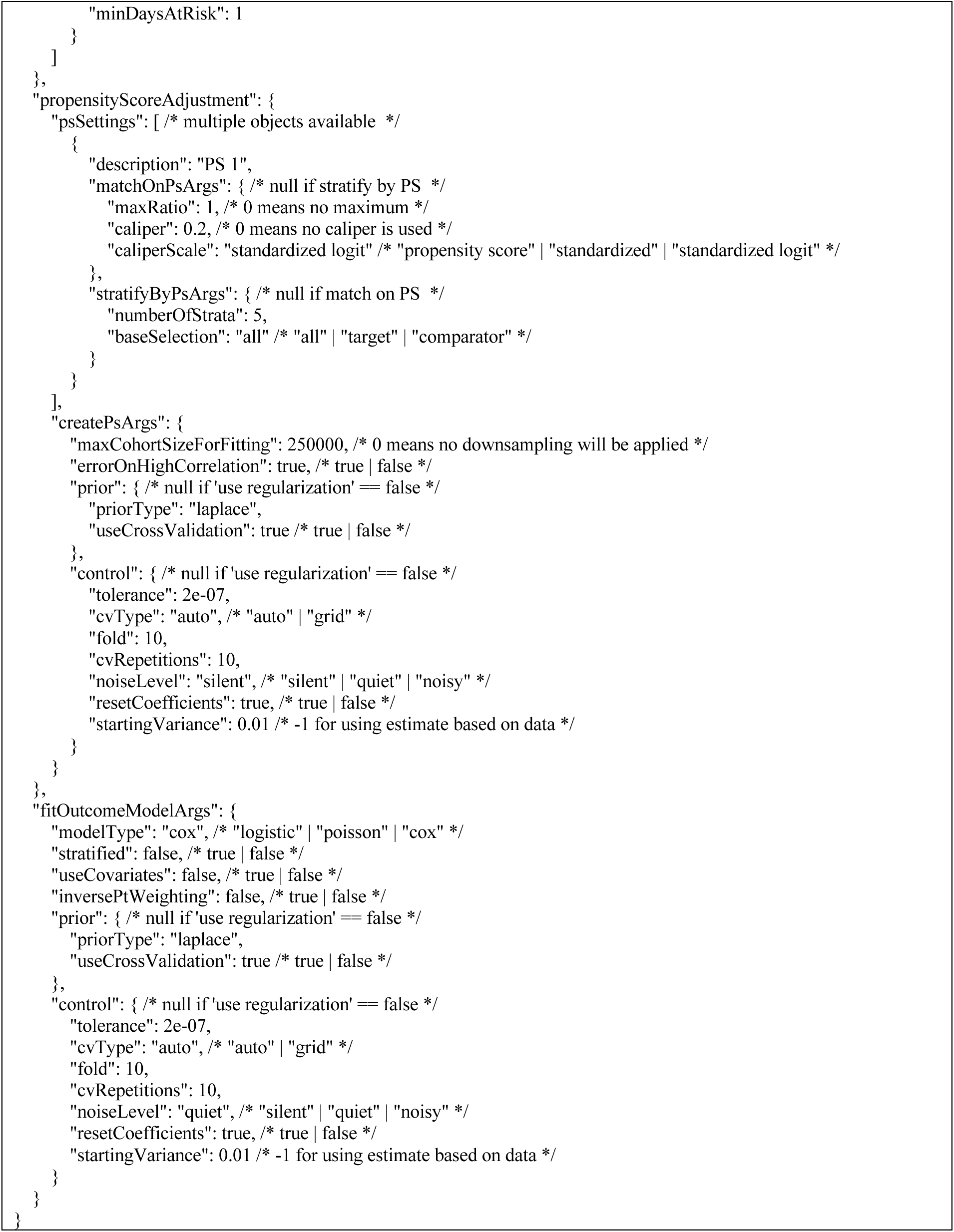
The predefined JSON template for the analysis specifications in the standardization step.

**Supplementary Material 2.**
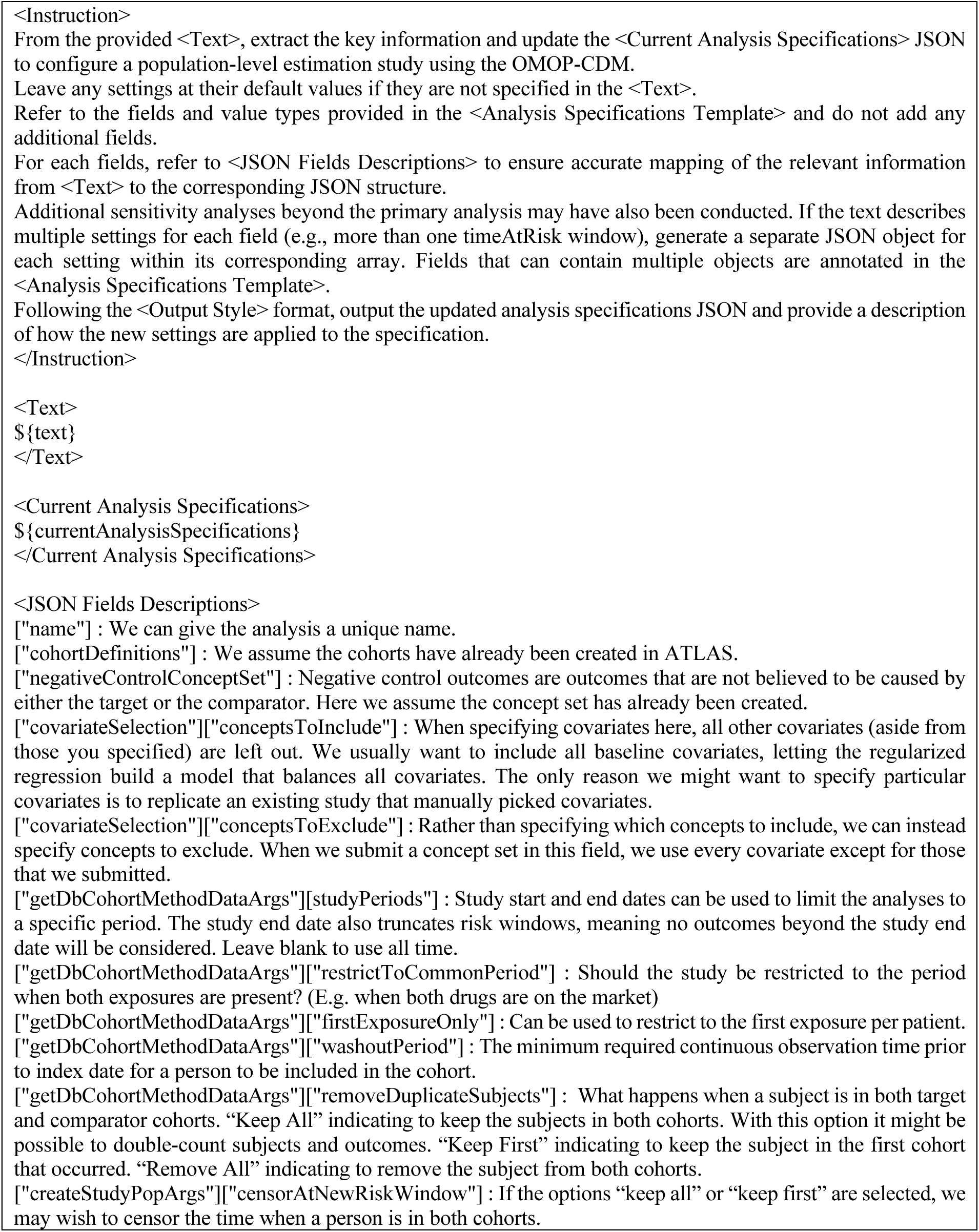
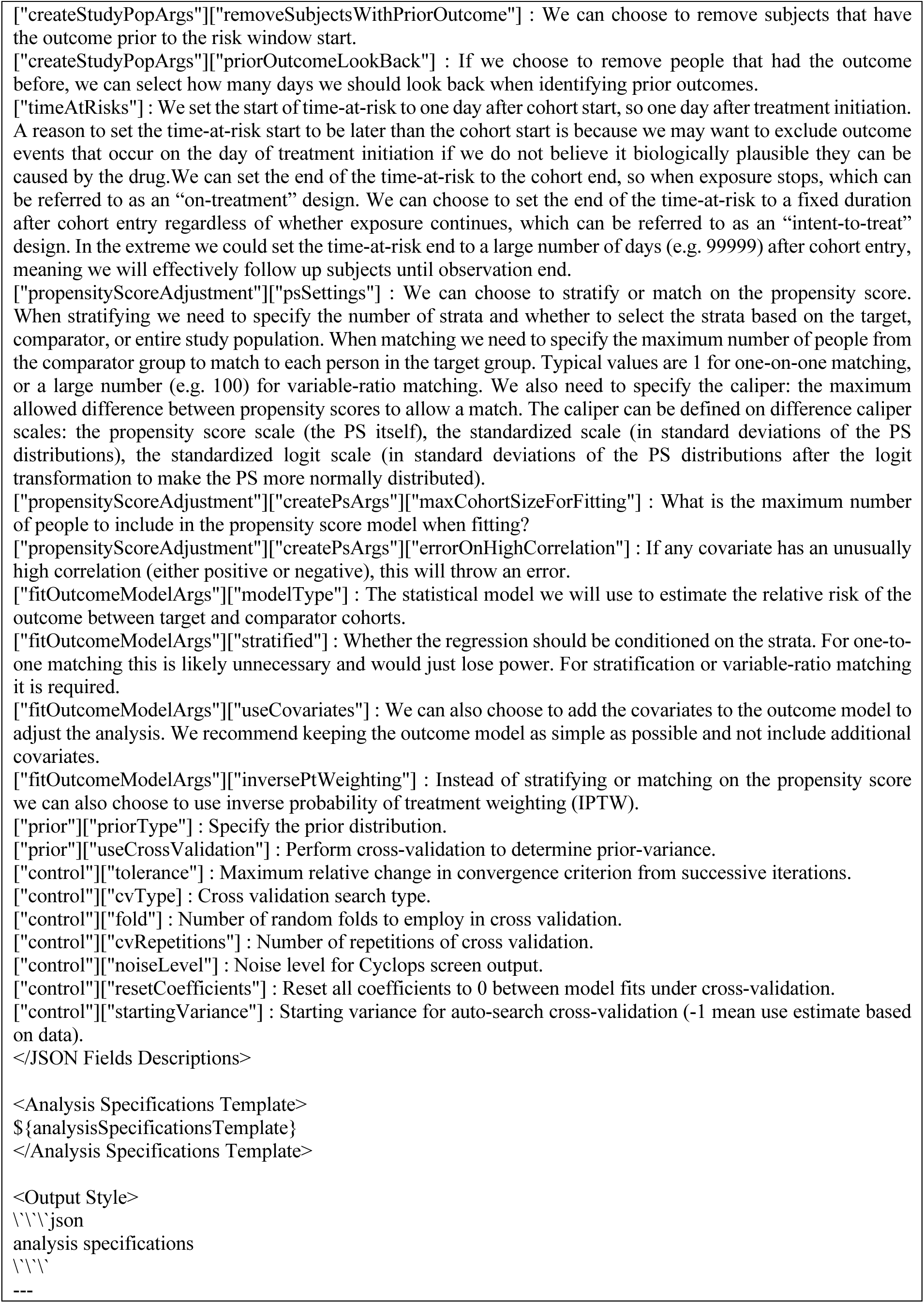
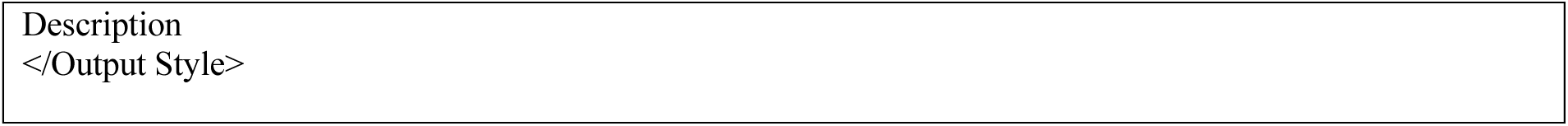
Prompt used for standardization step in THESEUS ${text} denotes the user’s study design described in natural language, ${currentAnalysisSpecifications} denotes the currently configured study design specifications, and ${analysisSpecificationsTemplate} denotes the predefined JSON template that constrains the structure of the generated analysis specifications.

**Supplementary Material 4.**
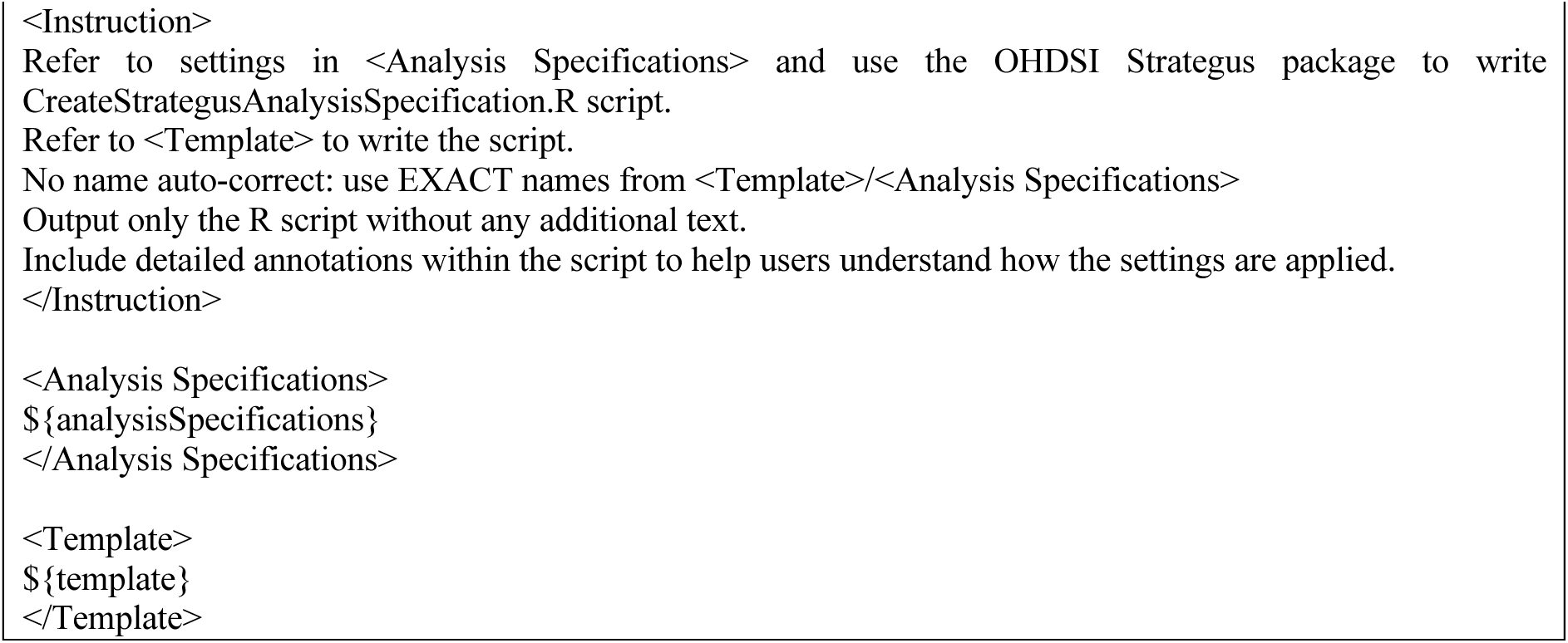
Prompt used for code generation step in THESEUS ${analysisSpecifications} denotes the current user-configured analysis specifications, and ${template} denotes the predefined Strategus R script template used to generate the CreateStrategusAnalysisSpecification.R.

**Supplementary Material 5.**
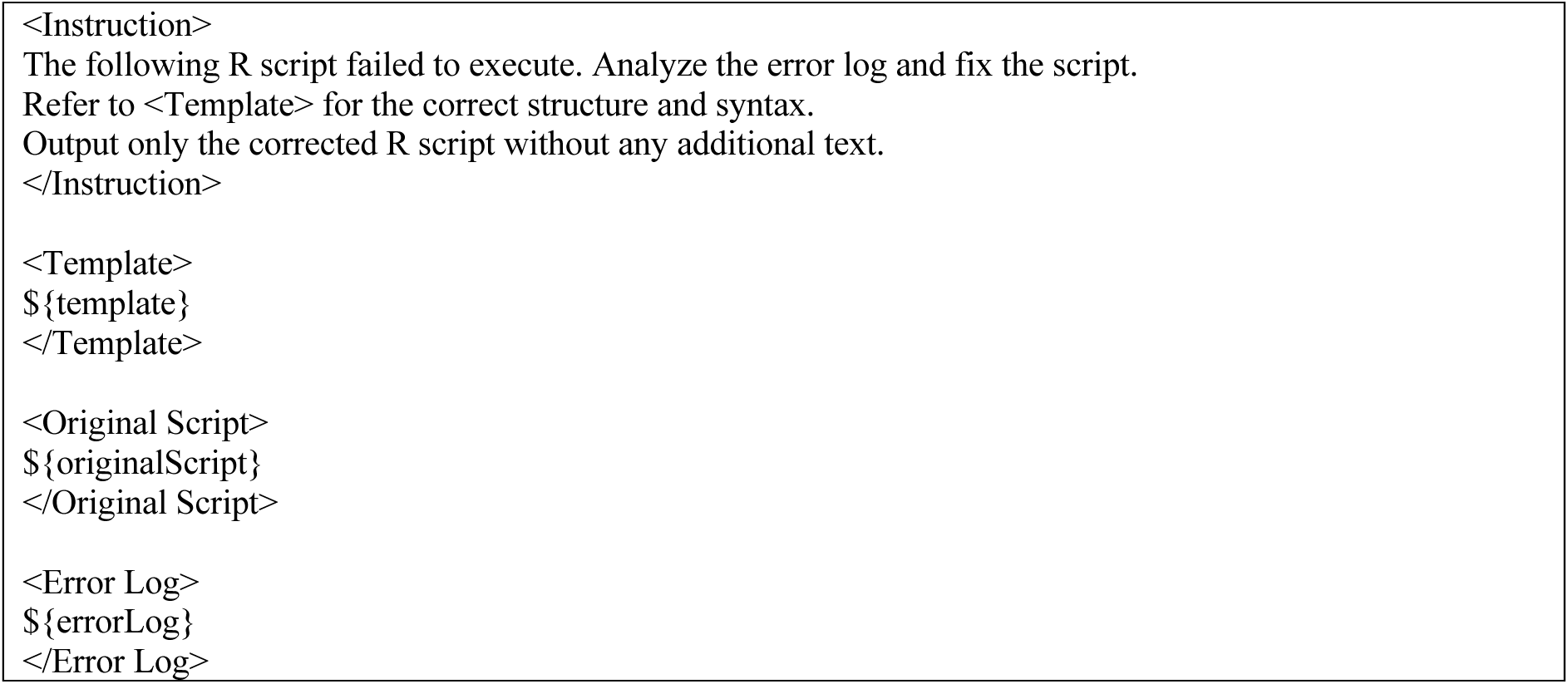
Prompt used for self-auditing step in THESEUS ${template} denotes the predefined Strategus R script template, ${originalScript} denotes the original R scripts that failed to execute, and ${errorLog} denotes the execution error log generated when running the original script.

**Supplementary Material 6.**
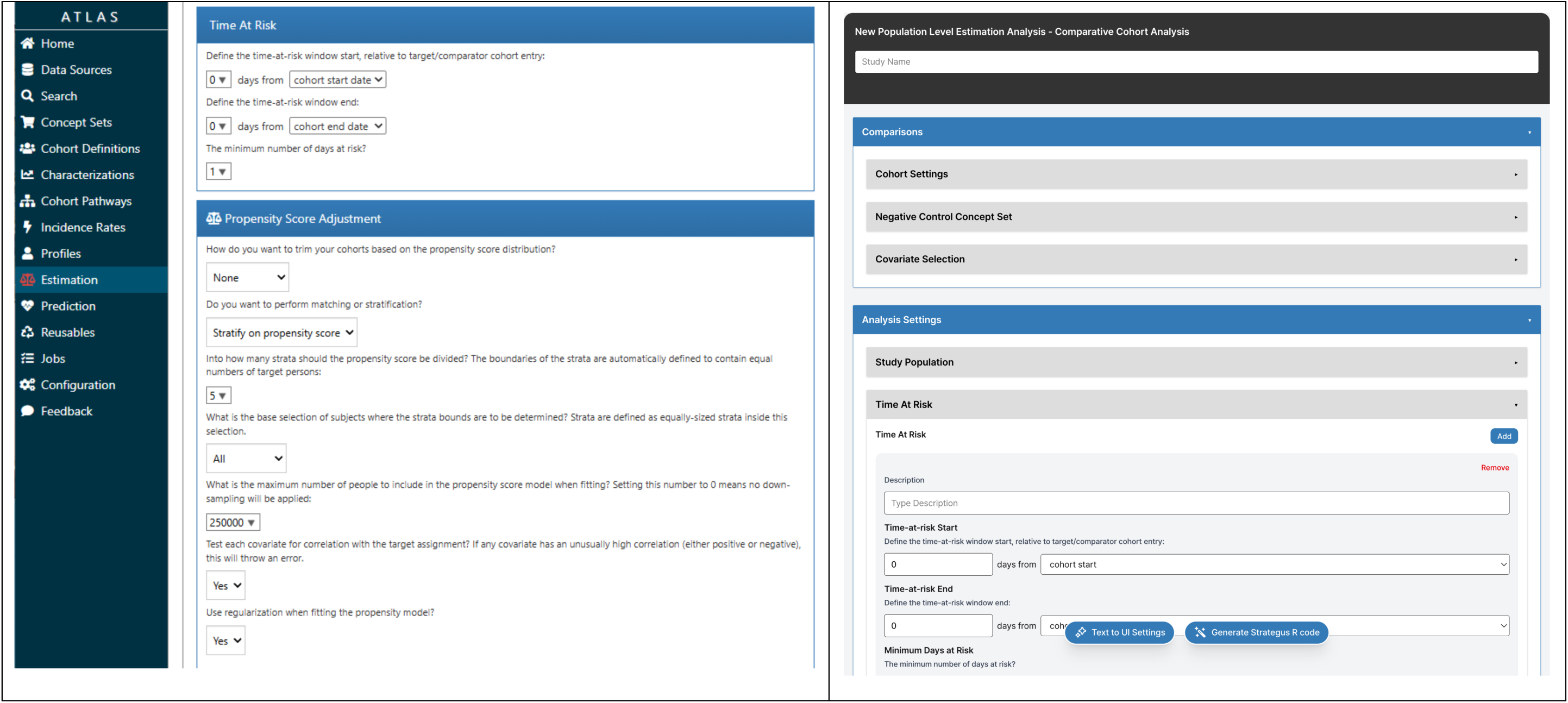
User interfaces of the original ATLAS (left) and THESEUS (right)

**Supplementary Material 7.** Large Language Model specifications.

| Vendor | Model | Model release date | knowledge cutoff |
| --- | --- | --- | --- |
| OpenAI | GPT-5.2 | 2025-12-11 | 2025-08 |
|  | GPT-4.1 | 2025-04-14 | 2024-06 |
| Gemini | Gemini-2.5-Pro | 2025-06-17 | 2025-01 |
|  | Gemini-2.5-Flash | 2025-06-17 | 2025-01 |
| Claude | Claude-Opus-4.5 | 2025-11-01 | 2025-05 |
|  | Claude-Haiku-4.5 | 2025-10-01 | 2025-02 |
| Deepseek | DeepSeek-V3.2 Thinking Mode | 2025-12-01 | unknown |
|  | DeepSeek-V3.2 Non-thinking Mode | 2025-12-01 | unknown |

